# Inferring global-scale temporal latent topics from news reports to predict public health interventions for COVID-19

**DOI:** 10.1101/2021.06.10.21257749

**Authors:** Zhi Wen, Guido Powell, Imane Chafi, David Buckeridge, Yue Li

## Abstract

The COVID-19 pandemic has highlighted the importance of non-pharmacological interventions (NPI) for controlling epidemics of emerging infectious diseases. Despite their importance, NPI have been monitored mainly through the manual efforts of volunteers. This approach hinders measurement of the NPI effectiveness and development of evidence to guide their use to control the global pandemic. We present EpiTopics, a machine learning approach to support automation of the NPI prediction and monitoring at both the document-level and country-level by mining the vast amount of unlabelled news reports on COVID-19. EpiTopics uses a 3-stage, transfer-learning algorithm to classify documents according to NPI categories, relying on topic modelling to support result interpretation. We identified 25 interpretable topics under 4 distinct and coherent COVID-related themes. Importantly, the use of these topics resulted in significant improvements over alternative automated methods in predicting the NPIs in labelled documents and in predicting country-level NPIs for 42 countries.

## 1 Introduction

It has long been understood that organized community efforts are required to control the spread of infectious diseases [1]. These efforts, called public health interventions, include social or non-pharmaceutical measures to limit the mobility and contacts of citizens and pharmaceutical interventions to prevent and limit the severity of infections. Non-pharmaceutical interventions (NPI) are not easily evaluated through experimental studies [2], so evidence regarding the effectiveness of these public health interventions is usually obtained through observational studies. This is also partly due to the lack of an efficient method to monitor NPI, which is challenging because their use is not recorded consistently within or across countries.

In the early stages of an emerging infectious disease, such as COVID-19, NPI tend to be particularly important due to the lack of specific pharmaceutical interventions. At the outset of the COVID-19 pandemic, recognizing the absence of systems for recording NPI, multiple groups initiated projects to track the use of interventions for COVID-19 around the world. These projects have relied on the manual efforts of volunteers to review digital documents accessible online with minimal coordination across projects [3]. This approach to monitoring interventions is not sustainable and is difficult to be implemented or extended to other infectious diseases [4].

Given that changes in the status of NPI are usually described in digital online media (e.g., government announcements, news media) (**Fig**. 1), machine learning methods have the potential to support the monitoring of NPI. However, the application of machine learning methods to this task is complicated by the need to generate interpretable results and also by the limitations in the amount and quality of labelled data.

**Figure 1:**
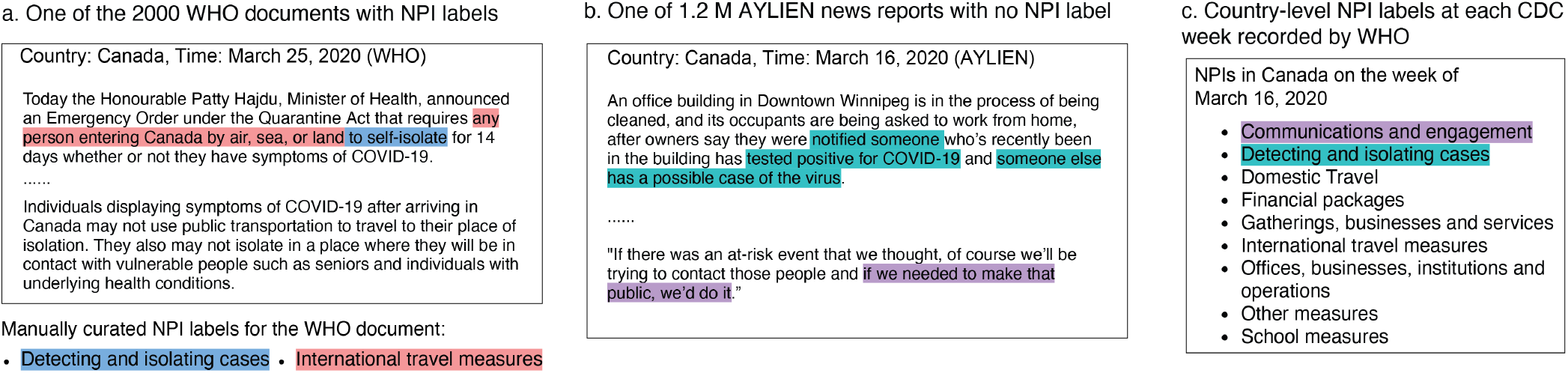
Examples of news reports and NPI labels. **a**. One of the 2000 WHO documents with two manually curated NPI labels. **b**. One of the 1.2 million AYLIEN news reports with no NPI label observed. Nonetheless, the sentences related to two NPIs are highlighted. **c**. Country-level NPI labels recorded at each CDC week. The observed NPIs in the same CDC week in Canada are highlighted, which are implicated in the AYLIEN news reports in panel b.

In this paper, we present a machine learning framework that uses online media reports to monitor changes in the status of NPIs that are used to contain the COVID-19 spread in 42 countries. Our method makes use of three types of data: around 2000 documents for which a group supported by WHO provided labels for 15 NPI, 1.2 million unlabelled news reports on COVID-19 made publicly available from a AI-powered news mining company AYLIEN, and country-level NPI labels at each CDC week (i.e., the epidemiological weeks defined by the Centers for Disease Control and Prevention) for 42 countries (**Fig**. 1). This type of event-based surveillance can inform situational awareness, support policy evaluation, and guide evidence-based decisions about the use of public health interventions to control the COVID-19 pandemic.

Our overarching goal in this work is to develop a topic modelling framework [5] to predict NPI labels at both the document-level and country-level to support the monitoring of public health interventions. To this end, we developed a three-stage framework called EpiTopics (**Fig**. 2). At the center of our approach is the use of probabilistic topic modelling, which facilitates the interpretation of our results and is well-suited to the noisy nature of online media. To address the limitation of the small number of NPI-labelled documents, we use transfer learning to infer the topic mixture from the labelled data using a topic model trained on a much larger amount of unlabelled COVID-19 news data. By classifying online media news into NPI categories based on interpretable, model-inferred topic distributions, we aim to automate many of the labour-intensive aspects of NPI tracking and enable systematic monitoring of interventions across larger geographical and conceptual scopes than are possible with currently used manual methods.

**Figure 2:**
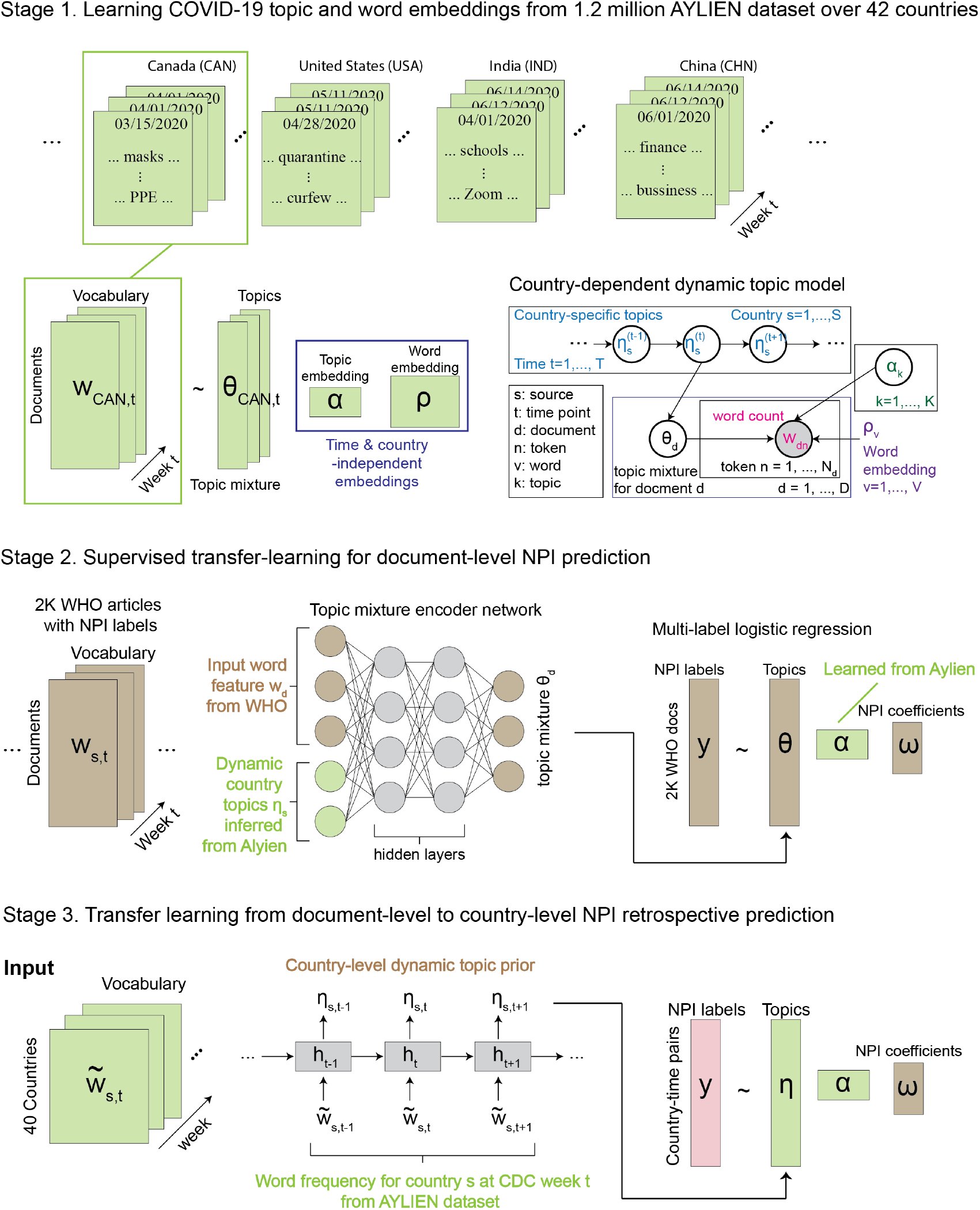
EpiTopics model overview. **a**. Unsupervised learning of COVID-19 topic and word embeddings from 1.2 million AYLIEN dataset over 42 countries. For each country, we extracted a set of news articles related to COVID-19 observed from November 1, 2019 to July 31, 2020 from AYLIEN. For each document (e.g., **w**_*CAN,t,d*_ for document *d* published at time *t* from Canada), we inferred their topic mixture (**θ**_*d*_). Meanwhile, we learned the global topic embedding (***α***) and word embedding (**ρ**) from the corpus. **b**. Supervised transfer-learning for document-level NPI prediction. We used the trained topic mixture encoder from Stage 1 to infer the topic mixture of the 2000 WHO news reports. The resulting topic mixture were then used as the input features to predict NPI labels in a logistic regression model. **c**. Supervised transfer-learning from document-level to country-level NPI prediction. Here we used country-specific topic trajectories ***η*** inferred at stage 1 as the input features to predict country-level NPI at each time point in a pretrained logistic regression model, where the linear coefficients were already fit at Stage 2 for the document-level NPI prediction task.

## 2 Results

### 2.1 EpiTopics model overview

EpiTopics is a machine learning framework for predicting non-pharmacological interventions (NPI) used to control the COVID-19 pandemic from the news data. The intended application of the framework is to predict each week, based on unlabelled news documents about COVID-19, any change in the status of multiple NPI at the country level. To this end, EpiTopics implements a three-stage machine learning strategy (**Fig**. 2), which is described in general terms below and in greater detail in the **Experimental Procedures** section.

### 2.2 Interpreting COVID-19 topics learned from the AYLIEN dataset

At the first stage, we sought to learn a set of unbiased latent topic distributions by mining a large corpus of news articles about COVID-19, but without the NPI labels. We used an open-access dataset of articles publicly released by an AI-news company AYLIEN, which included 1.2 million news articles about COVID-19 from 42 countries recorded from November 1, 2019 to July 31, 2020 (**Section** 4.1). All articles were related to COVID-19, but not necessarily related to particular NPI implementations. Although NPI labels were not available for these articles, in contrast to the smaller datasets with NPI labels (described next), this corpus was suitable for learning semantically diverse topics related to COVID-19 due to its size and coverage. To capture country-dependent topic dynamics, we developed an unsupervised model called MixMedia [6], which was adapted from the dynamic embedding topic model (DETM) [7] (**Fig**. 2a; **Experimental Procedures, Discussion**).

We experimented with different numbers of topics and chose 25 topics based on a common metric called topic quality [7] (**Experimental Procedures**; **Supplemental Information** S1; **Supplementary Fig**. S1). We annotated the 25 topics learned from the AYLIEN data at Stage 1 based on the most probable words under each topic (**Fig**. 3) and then grouped these topics into four themes: (1) central health-related issues; (2) broader social impacts; (3) specific locations; and, (4) indirectly related issues.

**Figure 3:**
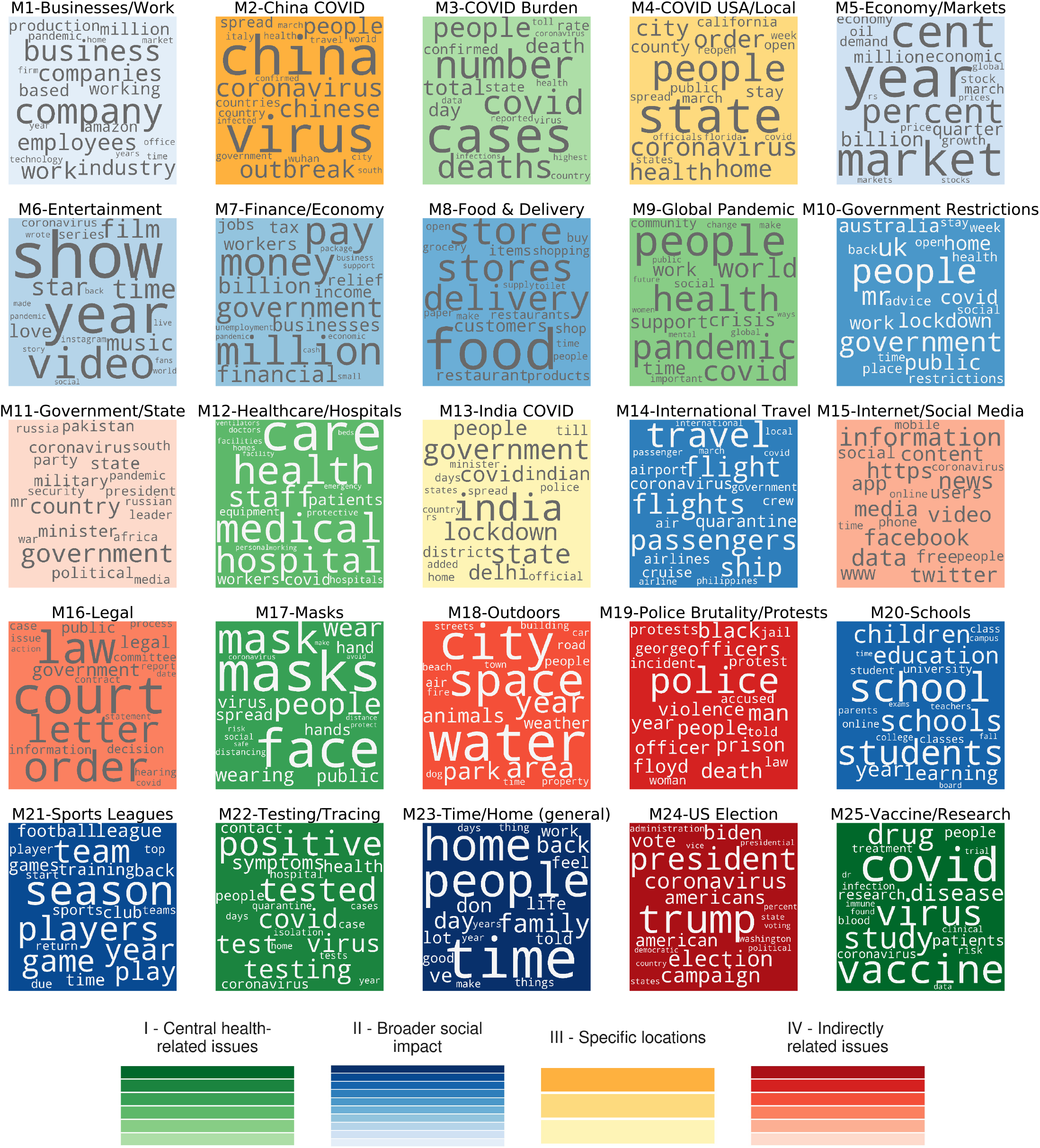
Learned topics and the top words under each topic. The sizes of the words are proportional to their topic probabilities. The background colors indicate the themes we gave to the topics.

Under Theme I, several topics addressed *central health-related issues* of the pandemic. For instance, the most frequent words in topic ‘M3 - COVID Burden’ included ‘deaths’ and ‘confirmed cases’; topic ‘M12 - Healthcare/Hospitals’ included ‘health’, ‘staff’, ‘doctors’, ‘beds’; topic ‘M22 - Testing/Tracing’ included ‘tested’, ‘positive’, ‘symptoms’; topic ‘M25 - Vaccine/Research’ included ‘vaccine’, ‘trial’, ‘drug’; topic ‘M17 - Masks’ included personal protective measures such as ‘wearing’, ‘masks’, (washing) ‘hands’, and ‘social distancing’; and topic ‘M9 - Global Pandemic’ included ‘health’, ‘pandemic’, and ‘world’.

Under Theme II, topics were associated with *broader social impacts*. For example, topic ‘M20 - Schools’ reflected changes in education including words such as ‘school’, ‘online’, ‘classes’, ‘exams’; topic ‘M14 - International Travel’ focused on changes in international travel, with top words like ‘flight’ and ‘passenger’; topic ‘M10-Government Restrictions’ included ‘lockdowns’ and ‘work’ (from) ‘home’; topic ‘M23 - Time/Home’ was related to spending time at home and family; topic ‘M8 - Food & Delivery’ discussed ‘restaurant’, ‘delivery’, ‘grocery’ and ‘stores’; topic ‘M21 - Sports Leagues’ was associated with sports teams and events; and topic ‘M6 - Entertainment’ was related to the impacts on entertainments such as ‘film’ and ‘music’. There were also topics related to financial impacts such as topic ‘M1 - Business/Work’ on ‘businesses’ and ‘companies’, topic ‘M5 - Economy/Markets’ on ‘stock markets’, and topic ‘M7 - Finance/Economy’ on impacts on ‘unemployment’, ‘income’, and ‘government financial packages’.

Under Theme III, some topics focus on impacts of the virus on *specific countries*. Topic ‘M2 - COVID China’ focuses on the pandemic in China and particularly in Wuhan city, topic ‘M4 - COVID USA/Local’ focused on the United States and its specific states and cities, and topic ‘M13 - COVID India’ focused on the pandemic in India.

Under Theme IV, a few topics were not directly related to COVID-19, although they were sometimes discussed in the context of COVID-19 as they reflected events occurring during the pandemic. For example, topic ‘M18 - Outdoors’ discusses ‘water’, ‘space’, and ‘parks’, which are activities with reduced risk of COVID-19; topic ‘M11 - Government/State’ was associated with states or governments; topic ‘M15 - Internet/Social Media’ focused on social media and the internet, which are crucial for distributing COVID-19 information; topic ‘M16 - Legal’ involves ‘court’, ‘law’, and ‘order’; topic ‘M19 - Police Brutality/Protests’ was associated with police brutality which sparked large-scale protests during the pandemic; and topic ‘M24 - US Election’ was associated with United States presidential election, for which the pandemic was a central issue.

Taken together, these observations indicate that the 25 topics inferred by our EpiTopics model from the AYLIEN news dataset represent diverse aspects of the COVID-19 pandemic. These inferred topics provide a rich foundation against which we can characterize online media reports about COVID-19. Notably, although subjective by nature, the above manual topic labelling is an important aspect of topic analysis, as the labels aid in the interpretation of the subsequent country-level analysis and NPI prediction tasks.

### 2.3 Country topic dynamics

Continuing to explore the results generated by EpiTopics at stage 1, we examined temporal patterns at the country level in the posterior distributions of topic popularity (***η***) inferred by our EpiTopics model. These patterns reflect changes over time in the COVID-19-related media news as the pandemic unfolded in each country (**Fig**. 4). As expected, media news in January and February from all of the 42 countries are predominantly about topic ‘M2 China COVID’. In March, the topic popularity began to diverge for different countries. Interestingly, the change in topic popularity for many countries meaningfully reflected the progression of the pandemic within those countries. For instance, topic ‘M13 India COVID’ started to rise at the end of March and stayed high for India media news. Asian countries, South Korea, China, and Japan tended to focus more on Topic ‘M12 China COVID’ even after February whereas the same topic dropped more sharply after February for countries in other global regions. Spain was the only country which exhibit high probability for topic ‘M21 SPORTS LEAGUES’, perhaps due to the strong impacts of COVID on these activities in Spain or possibly reflecting sampling bias from the data, which was focused on English-language media. In contrast to other countries, we observed more dramatic changes in topic dynamics for the United States with the focus shifting from topic ‘M2 China COVID’ in February to topic M4 for COVID in local regions of the US, then to multiple topics around June including topic M9 for the global pandemic, topic M24 US election, and the more general topic M23 for staying home. We caution that this theme may reflect the greater amount of articles collected for the United States compared to other countries. More generally, given that we considered only articles published in English, our results may be biased towards media content from English-speaking countries.

**Figure 4:**
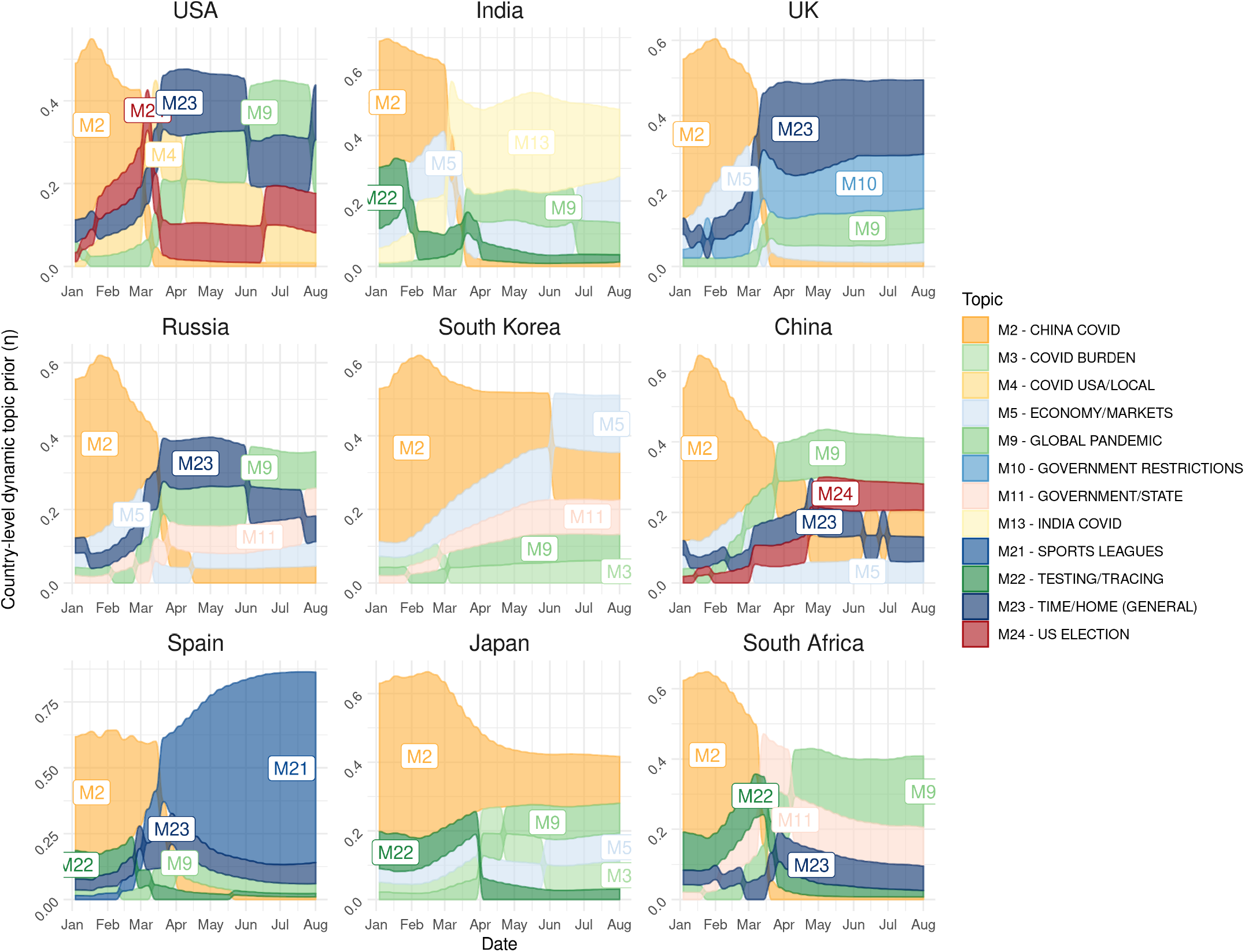
Temporal topics progression in example countries from January 2020 to July 2020. Different topics are represented in different colors. The height of each color block is proportional to the topic probability, in that country at that specific time. The topics were ranked in descending order vertically. To avoid cluttering the plot, only the top 5 topics were displayed for each country.

### 2.4 Predicting document-level interventions

At the second stage, we trained a linear classifier to predict the 15 NPI labels mentioned in the 2,049 news articles from the WHO dataset (**Section** 4.1) using the topic embeddings that were generated from the network encoder of the MixMedia model trained at Stage 1 (**Fig**. 2b; **Section** 4.4). We compared the prediction performance of EpiTopics and two baseline methds using the macro area under the precision-recall curve (AUPRC) and the weighted AUPRC **Table** 1). The baseline methods predicted NPIs from the bag-of-words (BOW) feature vector of a document (i.e., the raw word count) using either a linear classifier or a two-layered feed-forward network. Overall, EpiTopics outperformed the two baseline methods by a large margin, highlighting the benefit of incorporating topic mixtures (**Fig**. 5a). While the prediction accuracy greatly varied across individual NPIs, we achieved the best prediction performance for “Financial Packages”, “School Measures”, and “International Travel Measures” (AUPRC *>* 0.6). In addition, the use of topic modelling makes the results from our method highly interpretable via the learned classifier weights, which quantify the associations between the 25 topics and the 15 NPIs (**Fig**. 5b). For example, we observed that topic ‘M20 Schools’ was strongly associated with the NPI ‘School measures’, topic ‘M5 Economy/Markets’ was associated with ‘Financial packages’, and topic ‘M21 Sports League’ was associated with ‘Gatherings, businesses and services’. Notably, although some topics such as M21 alone did not have conceptual association with any NPI directly, their change correlated with changes in NPIs. They therefore could contribute to the prediction of NPIs. This result highlights the benefits of pre-training EpiTopics on a large unlabelled corpus to capture topics correlated with NPI.

**Table 1:** Area under the precision-recall curve (AUPRC) scores for document-level NPI prediction. The AUPRC scores are computed on individual NPIs, and then averaged without weighting (macro AUPRC) or weighted by NPIs’ prevalence (weighted AUPRC). Both BOW+linear and BOW+feed-forward use the normalized word vector (i.e., bag of words or BOW) for each document to predict NPI label. All methods are each repeated 100 times with different random seeds. Values in the brackets are standard deviations over the 100 experiments.

|  | **Weighted AUPRC** | **Macro AUPRC** |
| --- | --- | --- |
| BOW + linear | 0.165 (0.001) | 0.108 (0.001) |
| BOW + feed-forward | 0.149 (0.022) | 0.099 (0.007) |
| Document topic mixtures + linear | 0.408 (0.001) | 0.316 (0.001) |

**Figure 5:**
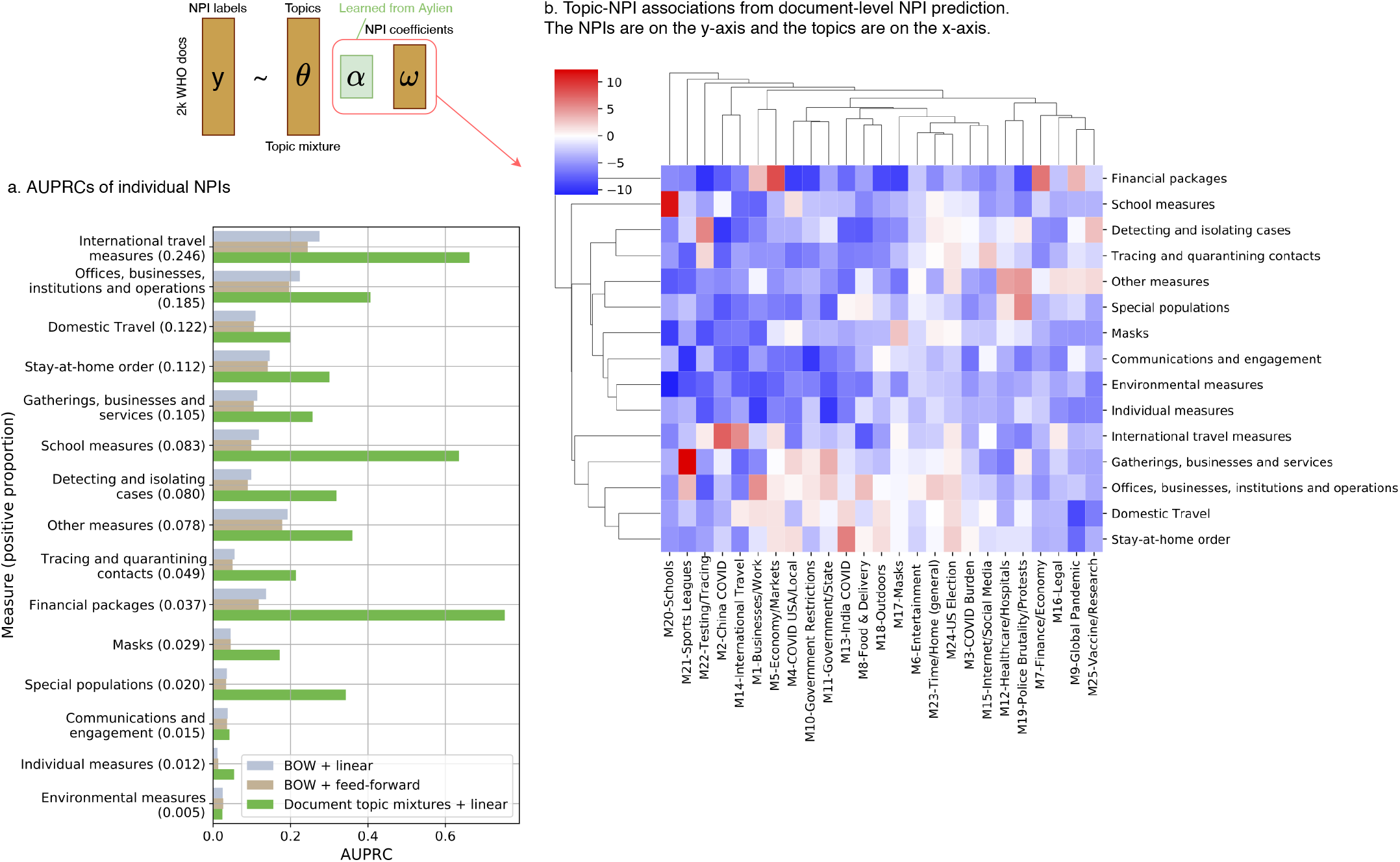
Document-level NPI prediction results. **a**. AUPRC scores on individual NPI predictions. We compared 3 methods: *BOW + linear* used bag of word (BOW) features (i.e., the word count vectors) to predict NPI with a linear classifier; *BOW + feed-forward* used a feed-forward neural network to predict NPI using the BOW features as inputs; *Document topic mixture + linear* : EpiTopics, which uses the inferred topic mixture from the trained encoder at Stage 1 to predict NPI by fitting a linear classification model. **b**. Topic-NPI associations learned by the linear classifier. The heatmap displays the linear coefficients with red, white, and blue indicating positive association, no association, and negative association, respectively.

### 2.5 Predicting country-level interventions

At the third stage, we performed transfer learning to predict the 15 NPI labels *at the country-level* for 42 countries by transferring model parameters learned from Stages 1 and 2 to Stage 3. The goal of the final stage is to predict whether there is *any change* in the status of a specific NPI at the country level from online media reports (**Fig**. 2c). Here a change in NPI status includes implementation, modification, and phase-out. We do not distinguish the types of change in our current model due to the limitations of the labelled data.

For input features, we used the ***α***-projected country-dependent topic posterior (***ηα***) inferred from the AYLIEN dataset at Stage 1. The weights (**ω**) learned at Stage 2 for the document-level NPI prediction were reused as the NPI-topic classifier weights. These weights were first used directly to predict NPIs at the country level without further training, an approach which we call *zero-shot* transfer. To improve upon this model, we also fine-tuned the topic-NPI coefficients **ω** to tailor the model towards country-level NPI prediction. We call this approach the *fine-tuned* model. Lastly, as a baseline model, we also trained a classifier by randomly initializing and fitting the topic-NPI coefficients **ω** while fixing the same ***α***-projected country-dependent topic posterior inferred from Stage 1. We call this approach the *from-scratch* model. Details were described in **Experimental Procedures**.

Overall, the ‘zero-shot-transfer’ model performed significantly better than ‘random’ model, and fine-tuning the transferred topic-NPI linear weights performed significantly better than training from scratch (**Fig**. 6c; **Table** 2). The results were mostly consistent for individual NPI prediction (**Fig**. 6a; **Fig**. S2). Furthermore, the predicted probabilities of NPIs were explained by the dominant country-level topic priors and their associations with each NPI (**Fig**. 7). For instance, the high probabilities for “Financial packages” in Philippines starting from March (**Fig**. 7), matched well with the true labels, and the high probabilities of this NPI were explained by the increasing probabilities of topics ‘M5 - Economy/Markets’ and ‘M7 - Finance/Economy’, and their strong association with NPI “Financial packages” learned in the classifier (**Fig**. 5b, **Fig**. 7).

**Table 2:** Area under the precision-recall curve (AUPRC) scores for country-level NPI prediction. Random baselines are each repeated 1000 times with different random seeds, and the rest are each repeated 100 times with different random seeds. Values in the brackets are standard deviations over the repeated experiments.

|  | **Weighted AUPRC** | **Macro AUPRC** |
| --- | --- | --- |
| Random eta, alpha and omega | 0.394 (0.007) | 0.307 (0.006) |
| Random eta and omega | 0.394 (0.007) | 0.307 (0.006) |
| Random omega | 0.398 (0.014) | 0.312 (0.011) |
| Zero-shot transfer | 0.427 (0.001) | 0.336 (0.001) |
| Trained from scratch | 0.446 (0.001) | 0.358 (0.001) |
| Fine-tuned | 0.488 (0.000) | 0.390 (0.000) |

**Figure 6:**
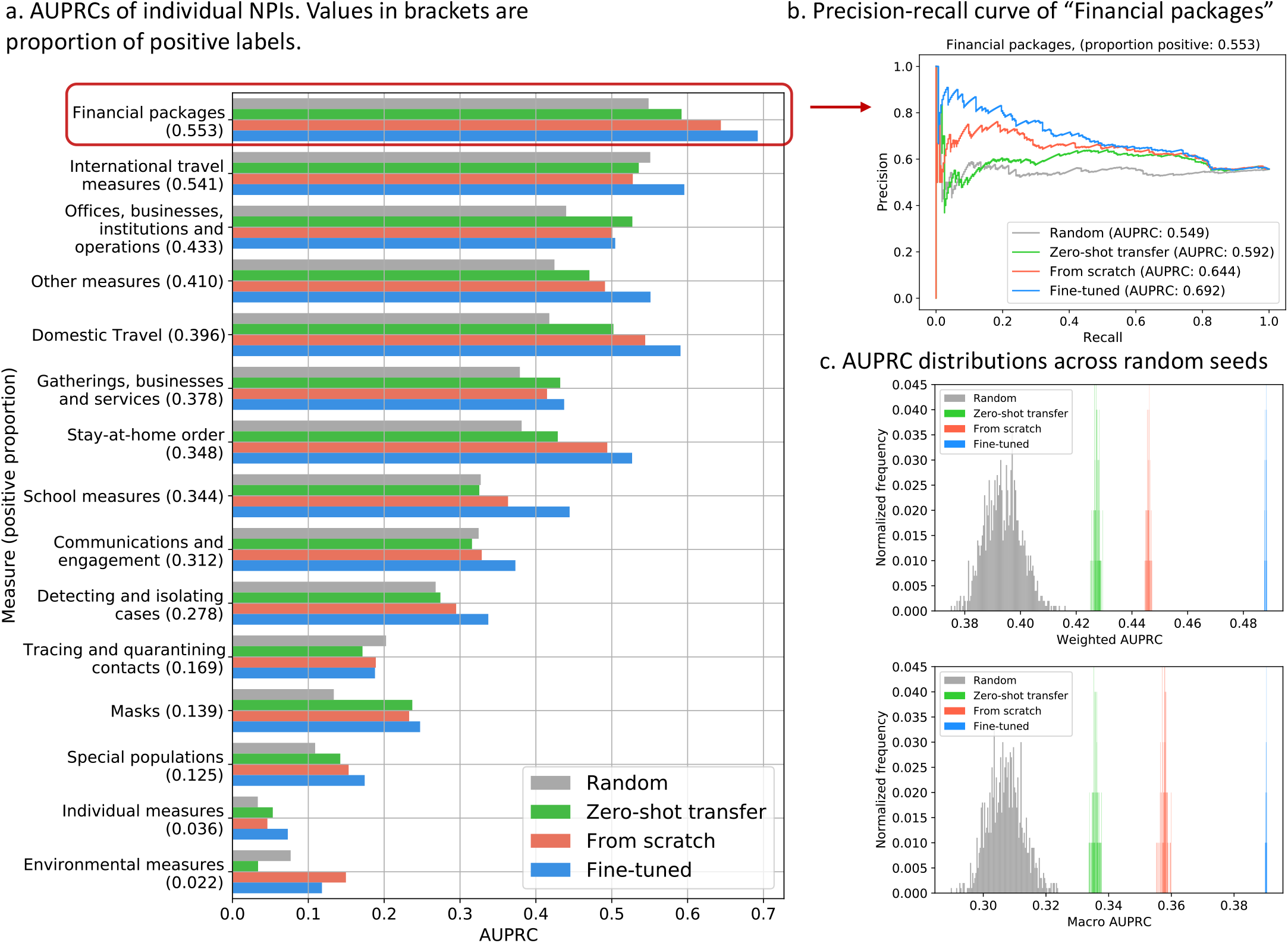
Country-level NPI predictions. **a**. AUPRC scores on individual NPI predictions at the country-level. ‘Random’, ‘Zero-shot transfer’, ‘From-scratch’, and ‘Fine-tuned’ are methods that predict NPI using random features, pre-trained linear coefficients from Stage 2 at the document-level NPI predictions, training the linear coefficients from random initialization, and fine-tuning the pre-trained linear coefficients from Stage 2, respectively. The numbers in the brackets indicate the fraction of the positive labels for that NPI, which are positively correlated with the corresponding AUPRC. **b**. Precision-recall curve of different methods on predicting “Financial packages”. **c**. Distributions of the weighted AUPRC scores and macro AUPRC scores over the 15 NPI predictions for the four methods across 100 repeated experiments with random initializations.

**Figure 7:**
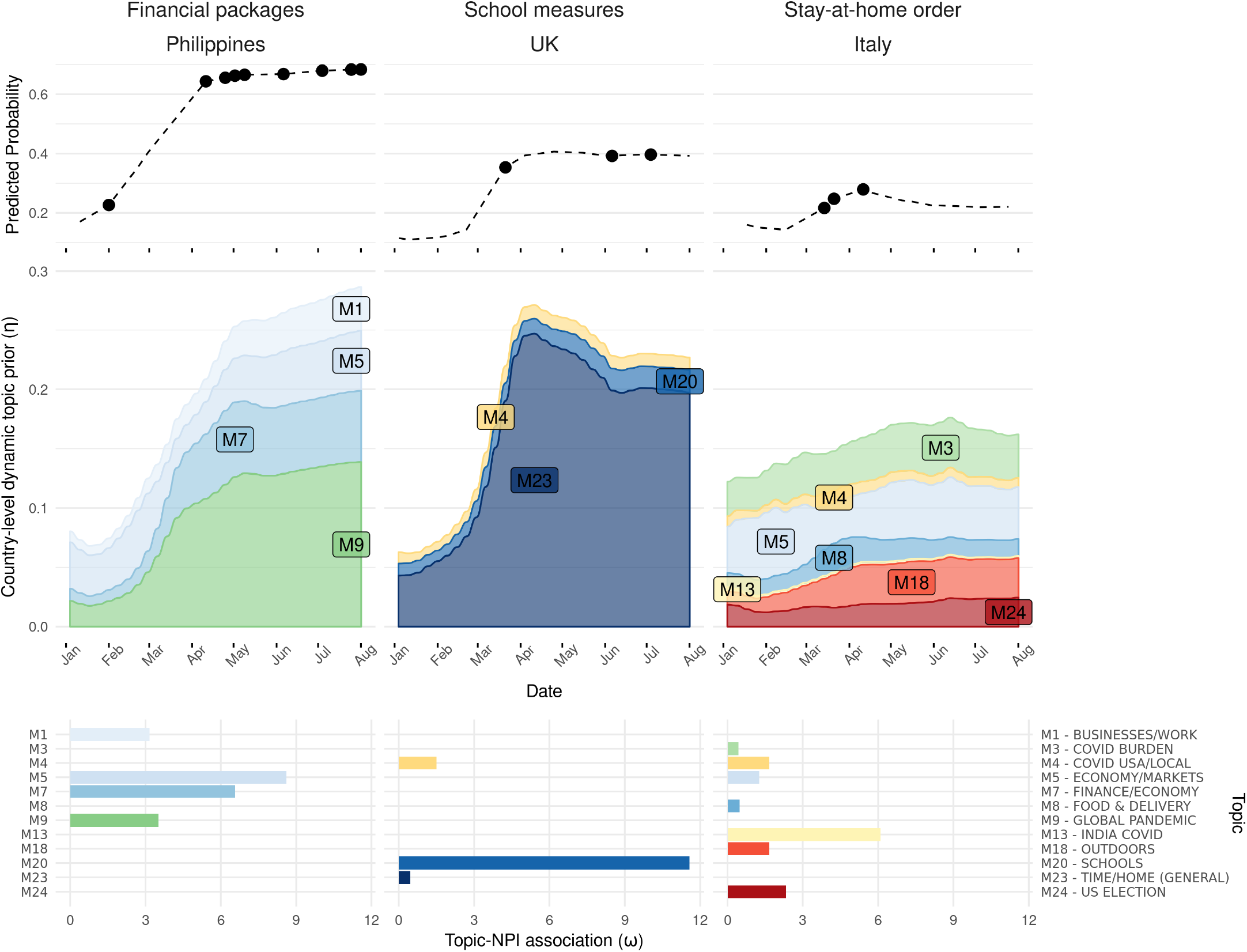
Example of topic dynamics and NPI predictions (using the *fine-tuned* method) for 3 select countries. In the top panel, the predicted probabilities over time of “Financial packages” in the Philippines, “School measures” in the UK and “Stay-at-home orders” in Italy are represented by the dashed lines, The dots represents the date of the NPI. These examples were selected to highlight a range of good to poor prediction accuracy (left to right). Predictions are informed by temporal topic dynamics within the AYLIEN dataset, represented in the middle panel. The bottom panel shows weights of each topic for a given NPI prediction, based on topic-NPI associations (**ω**) learned via predicting NPIs from the media articles in the WHO dataset. Topics with the predictive associations below 0 (i.e., *ω*_*k*_ *<* 0) are not shown.

We observed that the prevalence of NPIs was highly correlated with individual NPIs’ AUPRC scores, especially for the random baseline (**Fig**. 6a and **Fig**. S4b). This result was expected as the precision of completely random predictions is approximately equal to the proportion of the positive labels [8]. For non-random predictions, on a small dataset such as ours, it is also reasonable that the prevalence plays an important role. In addition to the prevalence of a NPI, the nature a NPI and the way it is defined and grouped can impact performance. In some groups, the NPIs can be diverse (e.g. “Other measures”), making these groups less conceptually coherent than other groups and harder to predict. Also, it is possible that some NPI status changes were not recorded by the trackers and therefore were missing in our dataset.

We also explored the feasibility of predicting country-level NPIs from *document-level topic mixtures* **θ**_*d*_ inferred from the AYLIEN dataset (**Section** 4.5). Compared with predicting from country-level topic priors (**Table** 3), we observed minor differences in performance of zero-shot transfer and fine-tuning. In addition, we trained another linear model from scratch using the document-level topic mixture to predict country-level labels associated with each document. While this approach has the advantage of using more documents to train a classifier, we observed similar performance compared to the fine-tuning approach. Notably, directly training on country-level topic mixture as input was much more efficient than training on the document-level topic mixture as there were 1 million documents and only a few hundred country-week pairs. Therefore, compared to the document-level alternative, the results highlight the advantage of directly operating on the inferred country-topic mixture in terms of both the interpretability and the computational efficiency.

**Table 3:** AUPRC scores for country-level NPI prediction from topics at document and country level. Values in the brackets are standard deviations. Random baselines are each repeated 1000 times with different random seeds, and the rest are each repeated 100 times with random seeds.

|  | **Topic level** | **Weighted AUPRC** | **Macro AUPRC** |
| --- | --- | --- | --- |
| Random eta, alpha and omega | Country | 0.411 (0.007) | 0.323 (0.006) |
| Random eta and omega | Country | 0.411 (0.007) | 0.323 (0.006) |
| Random omega | Country | 0.413 (0.017) | 0.326 (0.013) |
| Zero-shot | Country | 0.451 (0.001) | 0.352 (0.001) |
|  | Document | 0.448 (0.001) | 0.350 (0.001) |
| From scratch | Country | 0.455 (0.001) | 0.367 (0.001) |
|  | Document | 0.506 (0.000) | 0.398 (0.000) |
| Fine-tuned | Country | 0.503 (0.000) | 0.404 (0.000) |
|  | Document | 0.506 (0.000) | 0.397 (0.000) |

## 3 Discussion

In this paper, we present EpiTopics, a framework for surveillance of public health interventions using online news reports. This framework makes use of transfer-learning to address the limited amount of labelled data and topic modelling to guide the model interpretation. Specifically, we used transfer learning in two places. First, the topic-mixture encoder network trained on the 1.2 million unlabelled news reports related to COVID-19 at Stage 1 was transferred to inferring the topic mixture of the 2000 NPI-labelled WHO documents at Stage 2 (**Fig**. 2b). Second, we used temporal country-specific topics generated by the recurrent neural network encoder from Stage 1 as input features and the already trained document-level NPI linear classifier to predict the country-level NPI at Stage 3 (**Fig**. 2c). In terms of topic-modelling, our approach learned 25 interpretable topics, over 4 distinct and coherent COVID-related themes, whose country-specific dynamics appear to reflect evolving events in 42 countries. These topics contributed to significant improvements over alternative automated methods in predicting NPI from documents. While country-level predictions of some interventions were less accurate, the topic-NPI associations drawn from the previous step improved performance over the baseline models.

In the broader context, our work contributes to both public health and machine learning. From a public health perspective, the EpiTopics framework demonstrates the feasibility of using machine learning methods to meaningfully support automation of the surveillance of public health interventions. In the context of the COVID-19 pandemic, our methods can be used to enhance and facilitate manual efforts by research teams tracking interventions. While automated methods for screening have been used by existing COVID-19 trackers [3, 9], their limited accuracy and interpretability suggests that a hybrid or semi-automated approach is needed. With interpretable country-level topic dynamics, our model offers high-level insights along with classification guideposts for analysts attempting to monitor tens of thousands of NPI events around the world. More generally, expanding our approach to new corpora could further the development of rigorous methods for more general surveillance of public health interventions, which are not usually subject to routine monitoring. Therefore, EpiTopics complements the current, mainly manual approaches to monitoring NPI and has the potential to be expanded to the surveillance of public health interventions in general.

From a machine learning perspective, in contrast to the existing approaches that require large amounts of labelled data, our approach exploits the rich information from the vast amount of unlabelled news reports. We accomplish this by adapting the Dynamic Embedded Topic Model (DETM) framework [7]. We made two important modifications to the original DETM so that it would be more suitable for predicting NPI changes. First, we inferred temporal, *country-dependent* topic probabilities to represent the evolving pandemic situation within each country. Second, in contrast to the original DTEM, we inferred a set of *static* topic embedding to improve interpretation. This allowed us to analyze each topic separately without keeping track of the evolution of each topic over time within small time intervals (i.e., CDC weeks). Taken together, these modifications to the DETM allowed us to analyze the country-dependent topic trajectories over time by leveraging a large set of highly interpretable global topic distributions.

Our work can be extended in several future directions. First, our study is focused on the time frame from the beginning of COVID-19 to July 2020. Application of the EpiTopics framework to data covering a wider time frame is possible, for example through the Fall of 2021 when many countries experienced a fourth wave. Since the beginning of the second wave, many interventions were implemented, and vaccines were approved and administered across the globe, which would likely affect media coverage and NPI prediction.

Second, this study was conducted using two datasets consisting of COVID-19-related news reports each of which has their own limitations. The AYLIEN dataset lacks clear documentation on its sampling process and only includes records from English-language sources. These records may not be representative of media discussion of interventions across all countries and in some cases English language reporting of one country may not be representative of its media as a whole. While the WHO data has a very broad geographical coverage, issues with the validity of URLs in this dataset limited the records available for labelled data. Admittedly, these limitations of data sources restrict the generalizability of EpiTopics, in that countries not included in the training data are considered out-of-distribution and the EpiTopics model is unable to predict and monitor NPI for these countries. We intend to improve the generalizability of EpiTopics by exploring other sources of data such as CoronaNet [3] and data from Global Public Health Intelligence Network (GPHIN) [10] which may cover a broader range of countries, languages, NPIs, tasks and documents [9, 11].

Third, the documents are represented as a bag-of-words (BOW) for topic modelling. This representation omits rare words and stop words that are infrequent and too frequent, respectively. The order of words in sentences is also ignored. Therefore, we may lose some semantic meaning of the documents, especially when a sentence contains negation (e.g., deaths are not high). To this end, we intend to explore more complex neural language models such as ELECTRA [12] and BERT [13] at the expense of computations and sophisticated techniques such as attention mechanisms for model interpretability [14]. Also, these neural language models operate at the sentence level, whereas our model operates at the document and country levels. Future work can explore ways of exploiting manually labelled sentences [9] and investigate their performance on the task of NPI prediction.

Fourth, we can predict future country-level NPI at the next time point based on the previous time points using the country-level topic trend and previous NPI states. This can be done in an auto-regressive framework. Furthermore, incorporating active case counts (i.e., the number of newly infected people) may also potentially improve NPI prediction. The challenge here is the scarce observations of positive NPI changes at the country-level over time. Having more granular sub-region-level NPI labels (e.g., cities instead of countries) spanning over longer chronology of the pandemic may help training these more sophisticated models.

Lastly, geolocation extraction is a challenging task [15]. We expect that some proportion of countries were misclassified in the construction of the AYLIEN media dataset. While the large size of the dataset prohibits exhaustive validation, a non-exhaustive manual inspection of a subset of documents confirmed that a significant majority of country assignments were correct. To reduce the impact of this misclassification on our results, we limited our analyses to the country level rather than to the sub-region level, where misclassification is more likely. We intend to expand geolocation extraction in a future study in order to develop regional predictions.

In conclusion, our current work lays the methodological foundation for providing automated support for global-scale surveillance of public health interventions. Our work will greatly facilitate the manual NPI labelling process by automatically prioritizing relevant documents from a large-scale corpus. We envision that our work will also inspire further research to transform the way that public health interventions are monitored with advanced machine learning approaches.

## 4 Experimental Procedures

### 4.1 Datasets

#### WHO

Supported by the World Health Organization and led by a team at the London School of Health and Tropical Medicine, the WHO Public Health and Social Measures (WHO-PHSM) dataset (https://www.who.int/emergencies/diseases/novel-coronavirus-2019/phsm) represents merged and harmonized data on COVID-19 public health and social measures combining databases from seven international trackers. These individual trackers used mostly similar methods of manually collecting media articles and official government reports on COVID-19 interventions. The database has de-duplicated, verified and standardized data into a unified taxonomy, geographic system and data format. After validating entries across multiple trackers, the WHO team resolved conflicting information, such as different dates or disagreements in enacted measures, and carried forward only information from the best ranked sources (e.g. government documents over media reports and social media). We selected English-language records annotated with URL links to static content unchanged since its release. These pages were scraped using the Beautiful Soup and Newspaper3k Python libraries. Of over 30,000 records available at the time of writing, 8,432 records had valid URLs for scraping, of which 2,049 represented unique documents (articles or reports) that aligned with the period and countries in the larger AYLIEN dataset.

To balance the need for fine-grained NPI classes and the need for sufficient data points for model development, the WHO taxonomy covering 44 different categories of NPIs has been regrouped into 15 interventions (**Table** S1). The selection of categories to group together was determined by their conceptual similarity as well as their frequency. The prevalence of the 15 NPIs at the document and country levels are shown in **Fig**. S4 in Appendix S3. We split the data into a training set and a testing set with a 80%-20% ratio and tuned the hyperparameters on the training set.

#### AYLIEN

While the WHO dataset contain expert-curated documents with NPI labels, the documents available were too few and not representative of general media content on which a surveillance systems may operate. Therefore, in addition to the WHO dataset, we also use another dataset created by a News Intelligence Platform called AYLIEN (https://aylien.com/resources/datasets/coronavirus-dataset). Although this dataset has no NPI label, it has a lot more COVID-19 related news reports. In particular, the AYLIEN dataset has 1.2 million news articles related to COVID-19, spanning from November 2019 to July 2020. The data was compiled and generated using a proprietary NLP platform established by the AYLIEN company. The data was provided in the form of a json file that can be directly imported into the Python environment as a Panda DataFrame. We split 50% of data points (country-time pairs) into a training set and the remaining half into a testing set. The data processing is discussed next.

### 4.2 Data Processing

We adopted the same data processing pipeline for both datasets. Specifically, we removed white spaces, special characters, non-English words, single-letter words, infrequent words (i.e., words appearing in fewer than 10 documents). We also removed the same stop words as in [16] as well as common non-English stop words “la”, “se”, “el”, “na”, “en”, “de”. For WHO documents, we obtained country information from the “SOURCE” field associated with each WHO document. For AYLIEN documents, we took the “country” sub-field under the “source” field for the country origin of the document. Additionally, the content of the news reports are in the “body” field, and we use the “published_at” for the publication date of the reports. After removing documents without empty source fields, we have 1.2 million documents in the AYLIEN dataset. To minimize the effect of noise in time stamps and also to reduce sparsity of the data, we reduced the temporal resolution to weeks by grouping the documents and the NPIs within the same CDC weeks (epidemiological weeks defined by the Centers for Disease Control and Prevention; https://wwwn.cdc.gov/nndss/downloads.html).

In total, there are 1,176,916 documents in the AYLIEN dataset, over 42 countries and spanning 38 CDC weeks from November 2019 to July 2020. After applying the same pre-processing procedure to WHO and discarding the documents whose country or source is not seen in AYLIEN, we have 2,049 unique documents in the WHO dataset with NPI labels. There are 109377 and 8012 unique words in AYLIEN and WHO respectively. Among them, 366 unique words in WHO are not present in AYLIEN. Manual inspection confirms these 366 words are non-lexical or non-English and were discarded.

### 4.3 EpiTopics-Stage 1: Unsupervised dynamic embedded topic

The first stage of EpiTopics (**Fig**. 2a) is built upon our previous model called MixMedia [6], which was adapted from the Embedded Topic Model (ETM) [16] and the Dynamic Embedded Topic Model (DETM) [7]. The data generative process of EpiTopics is as follows:

1. Draw a topic proportion **θ**_*d*_ for a document *d* from logistic normal 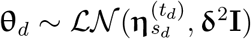:

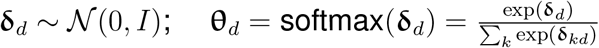

where *s*_*d*_ and *t*_*d*_ are indices for the country of the document *d* and the time at which document *d* was published, respectively;
2. For each token *n* in the document, draw word:

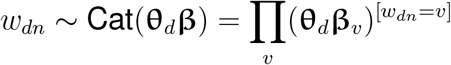

where **β** is a normalized *K* × *V* probability matrix with each row endowing a topic distribution over the vocabulary of size *V* :

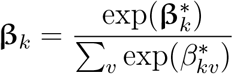

We further factorize the unnormalized topic distribution **β**^*\**^ into a *K* × *L* topic embedding matrix ***α*** and a *L* × *V* word embedding matrix **ρ**:

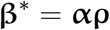

The above *K*-dimensional time-varying topic prior 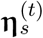 is a dynamic Gaussian variable, which depends on the topic prior at the previous time point of the same source:

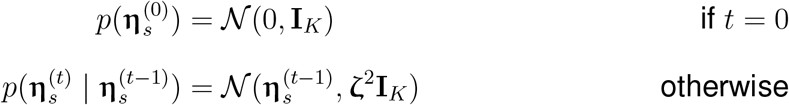

For the ease of interpretation, we assume that the *L*-dimensional topic embedding ***α***_*k*_ for each topic *k* is time-invariant and follows a standard Gaussian distribution [6]:

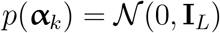

The word embedding **ρ** are fixed.

#### Inference

Our model has several latent variables including the dynamic topic prior ***η***_*s*_ per source *s*, the topic mixture per document **θ**_*d*_, the topic assignment per word per document token *z*_*dn*_, and the topic embedding ***α***_*k*_ per topic *k*. Word embedding **ρ** is treated as fixed point estimates and optimized via empirical Bayes. The data likelihood follows a categorical distribution:

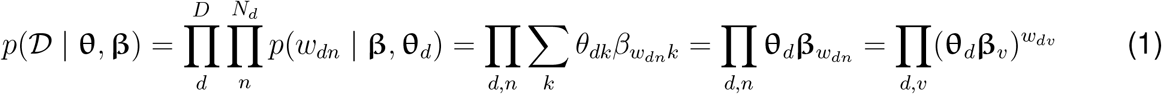

where 𝒟 = {**w**_1_, **w**_2_, · · ·, **w**_*D*_} denote the corpus, **w**_*d*_ denotes the frequency of each word in document *d, D* is the number of documents in the corpus, *N*_*d*_ is the number of tokens in document *d*, **β**_*kv*_ = softmax(***α***_*k*_**ρ**_*v*_) for word *w*_*dn*_ = *v* and topic *k*, and *w*_*dv*_ is the total count of word index *v* in document *d*. Therefore, the log likelihood is: log *p*(𝒟 | **θ, β**) =∑ _*d,v*_ *w*_*dv*_ log(**θ**_*d*_**β**_*v*_)

The posterior distribution of the other latent variables *p*(***η, α*, θ** | 𝒟) are intractable. To approximate them, we took an amortized variational inference approach using a family of proposed distributions *q*(***η, α*, θ**) [7]:

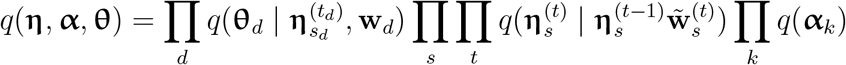

where

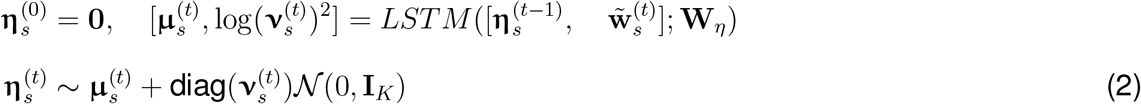

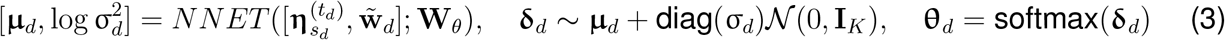

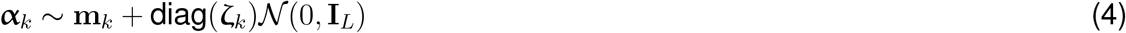

Here using the bag-of-word representation, **w**_**d**_ denotes a *V* × 1 vector of the word frequency of document *d* over the vocabulary of size 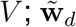 is the normalized word frequencies; 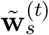 denotes average word frequency at time *t* for source *s*.

Using the variational autoencoder framework [17], the function *NNET* (**x**; **W**_***θ***_) is a feed-forward neural network parametrized by **W**_***θ***_; *LSTM* (**x**; **W**_*η*_) is a Long Short Term Memory (LSTM) network [18] parametried by **W**_*η*_. Because of the Gaussian properties, we use the re-parameterization trick [17] to stochastically sample the latent variable 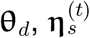, and ***α***_*k*_ from the their means with added Gaussian noise weighted by their variances as shown in Equations (2), (3), and (4), respectively.

To learn the above variational parameters Θ_*q*_ = {**W**_***θ***_, **W**_*η*_, **m**_*α*_, ***ζ***_*α*_}, we optimize the evidence lower bound (ELBO), which is equivalent to minimizing the Kullback-Leibler (KL) divergence between the true posterior and the proposed distribution *KL*(*q*(Θ)|*p*(Θ|𝒟)):

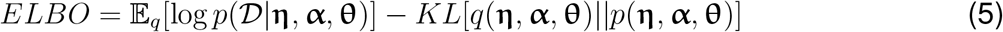

We optimize ELBO with respect to the variational parameters using amortized variational inference [7,17,19,20]. Specifically, we sample the latent variable ***η*** using Eq (2), **θ** using Eq (3), ***α*** using Eq (4) based on a minibatch of data. We then use those samples as the noisy estimates of their variational expectations for the ELBO (5). The ELBO is optimized with backpropagation using Adam [21].

#### Selecting the topic number based on topic quality

We use topic quality to select the best number of topics. Topic quality is calculated as the product of topic diversity and topic coherence. Topic diversity is defined as the percentage of unique words in the top 25 words from each topic across all topics, and topic coherence is defined as the average point-wise mutual information of the top-10 most likely words under each topic:

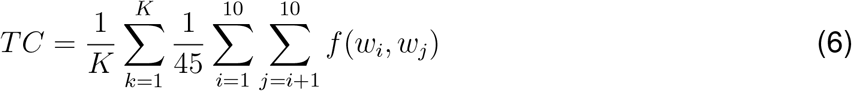

#### Implementation

EpiTopics-Stage1 or unsupervised MixMedia [6] is implemented and trained with PyTorch 1.5.0. We used 300-dimensional word and topic embeddings. We implemented the neural network in Eq (3) with a two-layer feed-forward network, with a hidden size of 800 and ReLU activation. We implemented the LSTM in Eq (2) with a 3-layer LSTM with a hidden size of 200 and a dropout rate of 0.1. We set the initial learning rate to 10^−3^ and reduced it by 1*/*4 up to the minimal value of 10^−7^ if the validation perplexity is not reduced within the last 10 epochs. The model was trained for 400 epochs with a batch size of 128. The number of topics 25 was chosen based on the best topic quality, as shown in **Fig**. S1.

### 4.4 EpiTopics-Stage 2: Predicting document-level interventions

At this stage, we use the unsupervised MixMedia trained at stage 1 to generate the expected topic mixtures of each article in the WHO dataset to predict the change in the status of an NPI reported in the article. The inputs to the classifier are the topic mixture of the WHO article produced by the MixMedia model trained on the AYLIEN data in the first stage. Specifically, the topic mixtures **θ**_*d*_ for WHO document *d* from country *s*_*d*_ observed at the 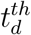 CDC week were produced from a feedforward encoder trained at stage 1:

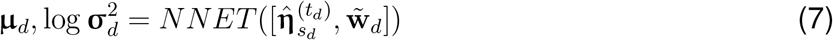

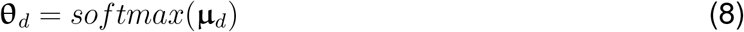

where *NNET* is the trained encoder of MixMedia in Eq (3), 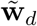 is the normalized word frequencies of document *d*, and 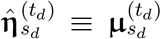 is the expectation of the country-level topic mixture for country *s*_*d*_ at time *t*_*d*_, which is produced by the trained LSTM of MixMedia in Eq (2).

We then fit a linear classifier to predict the 15 NPIs for document *d* using its topic mixture **θ**_*d*_:

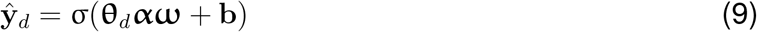

where σ(*x*) = 1*/*(1 + *e*^−*x*^) is the sigmoid function, ŷ_*d*_ is the vector of predicted probabilities of the 15 NPIs, **ω** is the linear weights and **b** is the bias vector. Notably, here we projected the topic mixture (**θ**) by the topic embedding (***α*** – also learned from Stage 1) to yield a document embedding input matrix. This ***α***-embedding allowed for higher dimensional representation of concepts relevant to NPI for better prediction performance than using **θ** alone. To predict the NPI mentioned in an article, we fit a logistic regression model. We chose a linear classifier here for the ease of interpretability and also due to small training sample size. The associations between topics and NPIs can be obtained by inspecting the inner product of the topic embedding and regression coefficients: ***αω***, which gives a *K* ×15 matrix indicating the associations between the K topics and 15 NPIs (**Fig**. 5).

The loss function here is cross-entropy plus the L2-regularization:

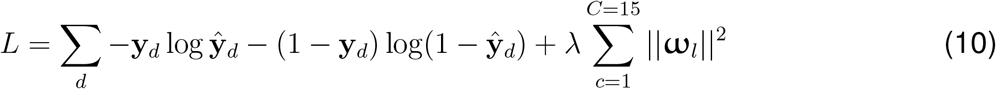

#### Implementation

The linear classifier was implemented with PyTorch 1.5.0. We used Adam optimizer with a learning rate of 6 × 10^−3^ and L2-penalty *λ* of 10^−5^. We trained the classifier for 400 epochs with a batch size of 512. To mitigate class imbalance over the 15 NPIs, we adjusted the weights for the per-NPI loss by up-weighting the loss of minority NPI class labels and down-weighting the loss of majority NPI class labels:

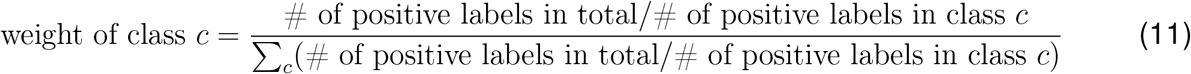

### 4.5 EpiTopics-Stage 3: predicting country-level interventions

#### Prediction from country-level topic priors

After the linear classifier for document-level NPI prediction is trained, its weights are transferred to the prediction task for country-level NPI labels:

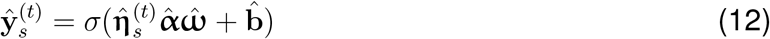

We experimented three models as follows.

For zero-shot-transfer model, we directly use the *country-level topic mixture* ***η*** and topic embedding ***α*** from learned Stage 1 and the trained linear classifier coefficients 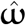 and biases 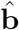 from Stage 2.

For the fine-tune model, the classifier weights are further updated for predicting country-level interventions. Specifically, we initialize the classifier weights with the learned parameters 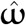 and further adjust it to predict country-level NPIs.

For the from-scratch model, we initialize the classifer weights by random values sampled from standard Normal and learn the weights from scratch to predict the 15 NPIs from the training country-time pairs. More details are described in Section 4.6.

#### Prediction from Document-level Topic Mixtures

We also experimented with an alternative approach by predicting country-level NPIs using *documents’ topic mixtures*. Specifically, the country-level NPI probabilities are calculated by taking the average of the predictions of the document-level NPI for the corresponding country and time:

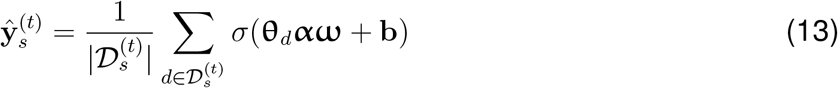

where 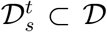 is the set of documents associated with country *s* and time point *t*. Therefore, this method can only be trained on predicting NPIs for the country and time pairs, where there is at least one document observed. Similarly to the method above, predictions can be made in a zero-shot transfer way, or the classifier weights are updated from the learned weights for the fine-tuned model or from random weights for the from-scratch models.

### 4.6 Experiments

#### Baselines

We omitted the comparison with other models at EpiTopics-Stage1 and refer the readers to our MixMedia work for comparing the topic quality with baseline topic models [6]. For document NPI predictions, we compared the EpiTopics-Stage2 model (i.e., using the inferred document-level topic mixtures of each WHO document to predict their NPI labels) against the baseline models that are trained to directly use the input word frequencies as the bag-of-word (BOW) to predict WHO labels. We experimented with two baseline methods namely linear (BOW + linear) and feed-forward network (BOW + feed-forward).

The BOW + linear baseline is trained with learning rate 8 × 10^−5^ and weight decay 0.002 for 100 epochs. The BOW + feed-forward baseline is trained with learning rate 10^−4^ and weight decay 0.002 for 100 epochs. The feed-forward network is implemented with two fully connected layers with a hidden size of 256 and ReLU activation. Fine-tuning used learning rate 6 × 10^−2^ and weight decay 1 × 10^−3^ with a batch size of 1024. The from-scratch model used learning rate 2 × 10^−3^ and weight decay of 2 × 10^−1^ with a batch size of 1024.

For country-level NPI prediction, we compared our proposed transfer-learning approach EpiTopics-Stage3 against two baseline models: (1) training from scratch; (2) model using randomly generated weights. For the random model, we randomly initialized the country-specific topic mixture 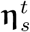, topic embedding ***α*** and/or classifier parameters {**ω, b**} and did not update their weights. To measure the effect of different levels of randomness, we experimented with three random baselines as follows:

- Random country-topic mixture ***η***, random topic embedding ***α***, and random topic-NPI regression coefficients **ω**: setting 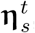, ***α*** and {**ω, b**} to random numbers
- Random ***η*** and **ω**: setting 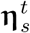, and {**ω, b**} to random numbers
- Random **ω**: setting {**ω, b**} to random numbers

Intuitively, the first random baseline preserves no information gained from the training of document-level NPI predictions, and thus represents a complete random model. The other two baselines preserve some level of information and thus may perform better than the first baseline.

#### Evaluation

Since NPI prediction is a multi-label classification problem, and that different NPIs can have different optimal thresholds, we used weighted AUPRC and macro AUPRC as evaluation metrics. We computed the Area Under the Precision-Recall Curve (AUPRC) for each NPI individually. We took either the unweighted average of them (macro AUPRC) or the weighted average of AUPRC by each classes’ prevalence (weighted AUPRC).

### 4.7 Our contributions in contrast to the existing computational methods

Other researchers have applied supervised learning to online media [3] and Wikipedia articles [9] to identify COVID-19 NPI. These approaches either entirely rely on the limited manually labelled data in a supervised learning framework or are difficult to interpret. In contrast, our approach has less demand for labelled data as it exploits large-scale unlabelled data via unsupervised and interpretable topic modelling combined with the transfer-learning strategy. Topic modelling has been used for surveillance of online media to detect epidemics [22–25] and to characterize news reports [26] and social media [27] in the context of COVID-19 or other outbreaks. Our work differs from these applications due to a focus on tracking NPI as opposed to detecting epidemics. By adopting a dynamic topic model, EpiTopics goes beyond the document-level to support inference at the country level.

## Data Availability

The datasets analyzed during the current study are from publicly available repositories or data portals. The acquisition and quality control steps for all datasets are included in the supplementary information.

https://github.com/li-lab-mcgill/covid-npi

## 5 Acknowledgements

This work is supported by CIHR through the Canadian 2019 Novel Coronavirus (COVID-19) Rapid Research Funding Opportunity (Round 1), (Application number: 440236).

## 6 Author contributions

Y.L. and D.B. conceived the study. Y.L. and Z.W. developed the model with critical helps from

D.B. and G.P.. I.C. collected and processed the data. Z.W. implemented the model and ran the experiments. Z.W., G.P., D.B., and Y.L. analyzed the results and wrote the paper.

## Supplemental information

### S1 Experiment Details

The number of topics 25 is chosen based on the best topic quality, as shown in **Fig**. S1.

**Figure S1:**
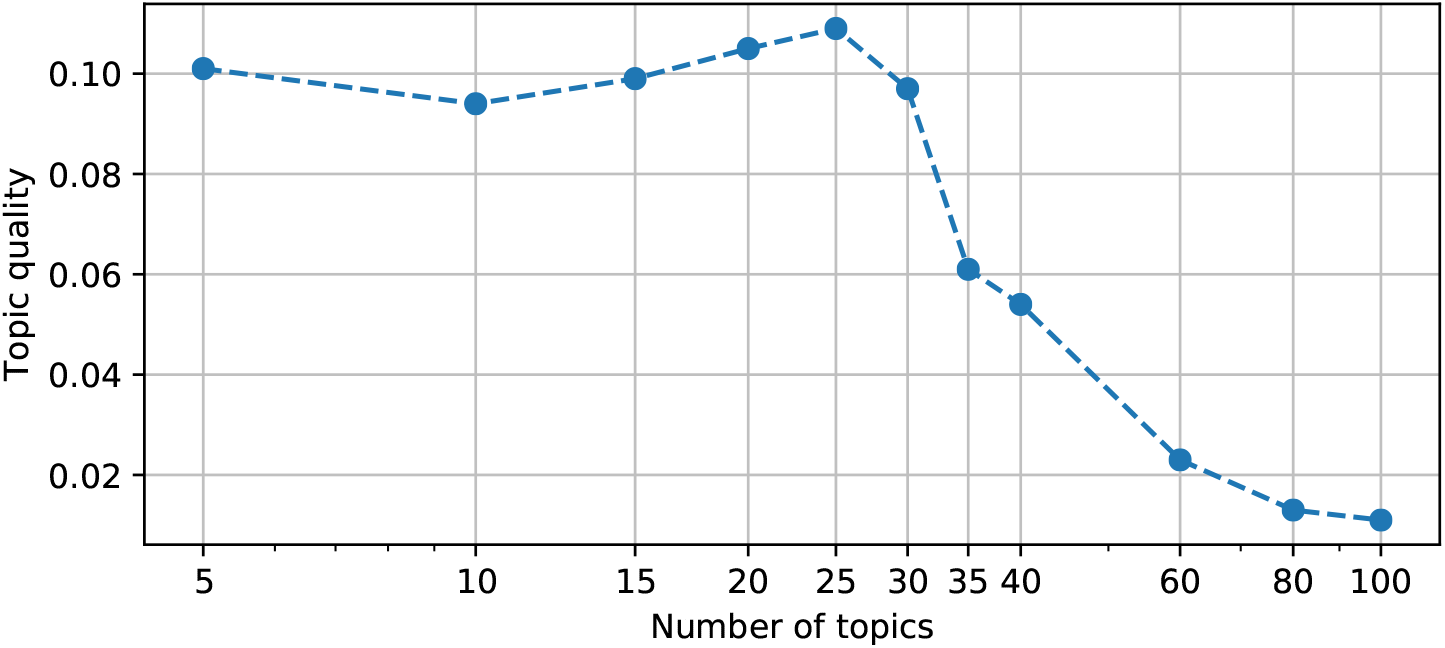
Topic quality versus the number of topics. Using 25 topics leads to the highest topic quality, and that number is used throughout this work. The x-axis is log-scaled.

### S2 Precision-Recall Curves of All NPIs

The precision-recall curves for the all NPIs are in **Fig**. S2.

**Figure S2:**
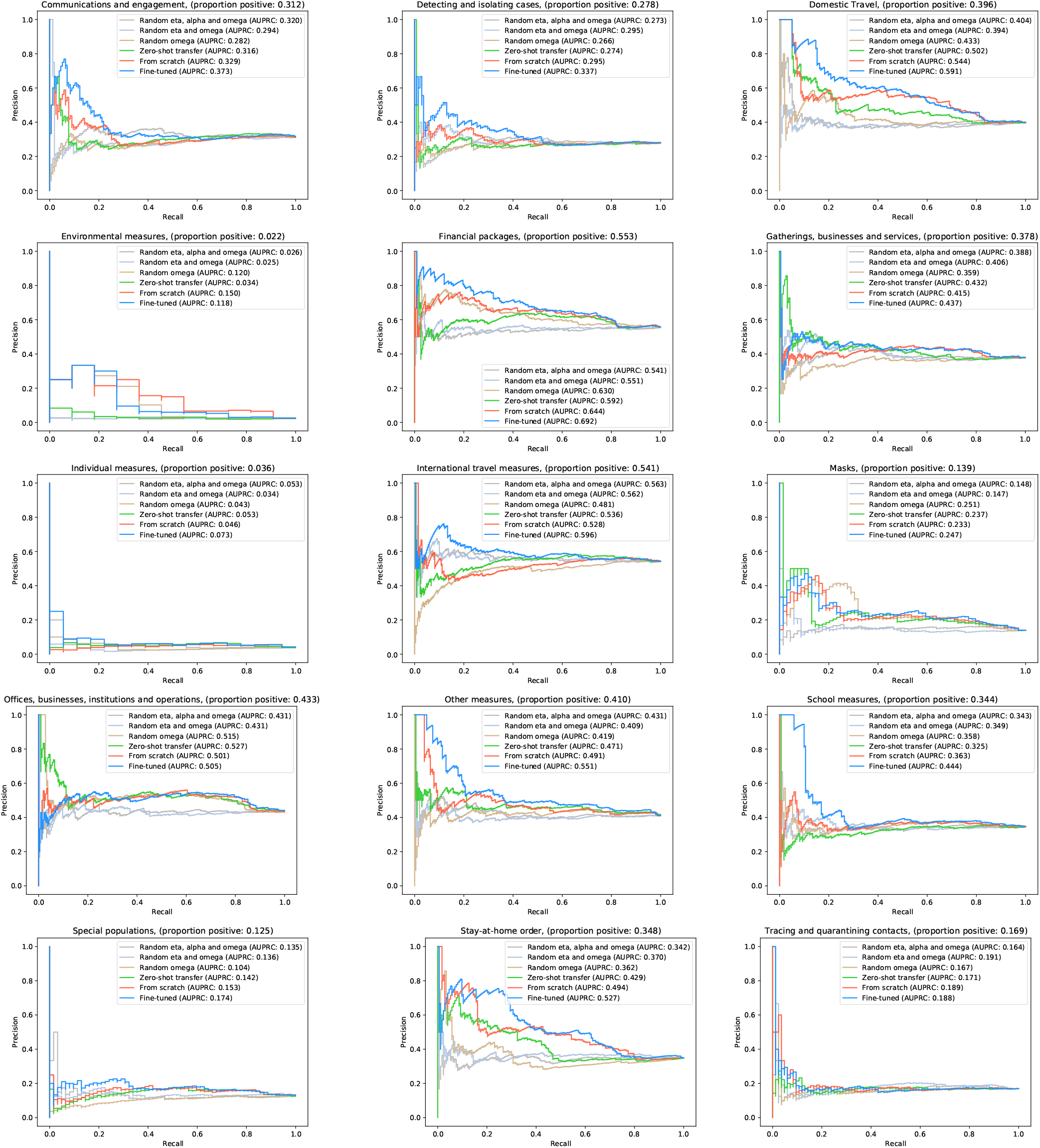
Country-level precision-recall curves of all 15 NPIs over 42 countries. Similar to **Fig**. 6b, each subfigure corresponds to one NPI, and together they correspond to all the NPIs in **Fig**. 6a. The predictions are made by a linear classifier on the country-specific topic mixture, and different methods in comparison learn the classifier weight differently as described in **Section** 4.5. In particular, Random, zero-shot transfer, from-scratch, and fine-tuned are methods that predict NPI using random features, pre-trained linear coefficients from Stage 2 on document-level NPI predictions, training the linear coefficients from random initialization, and fine-tuning the pre-trained linear coefficients from Stage 2, respectively. The numbers in the brackets indicate the proportion of the positive labels for that NPI, which are positively correlated with the corresponding AUPRC.

### S3 Dataset Details

The number of positive labels at the document-level in the WHO dataset and at the country-level are shown in **Fig**. S4.

**Figure S3:**
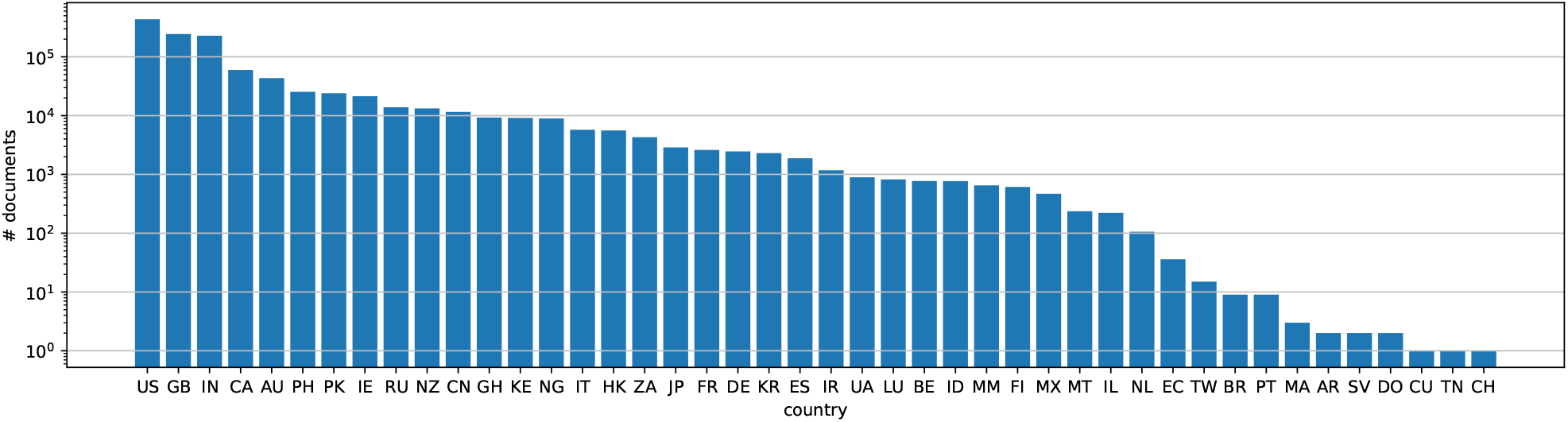
Number of documents of each country in the AYLIEN dataset. The y-axis is log-scaled.

**Figure S4:**
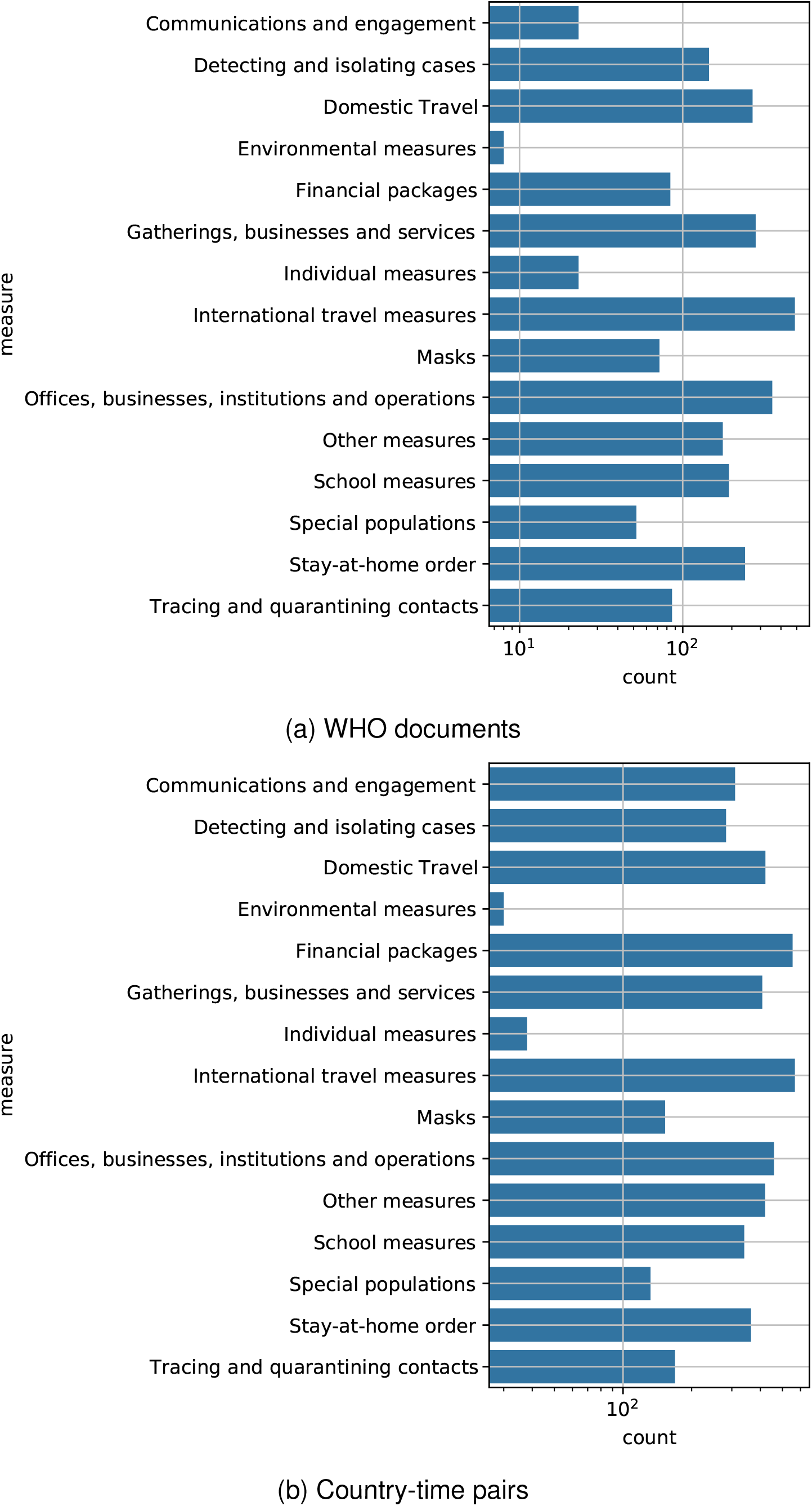
The number of positive labels at the document and country levels. The x-axis is log-scaled.

**Figure S5:**
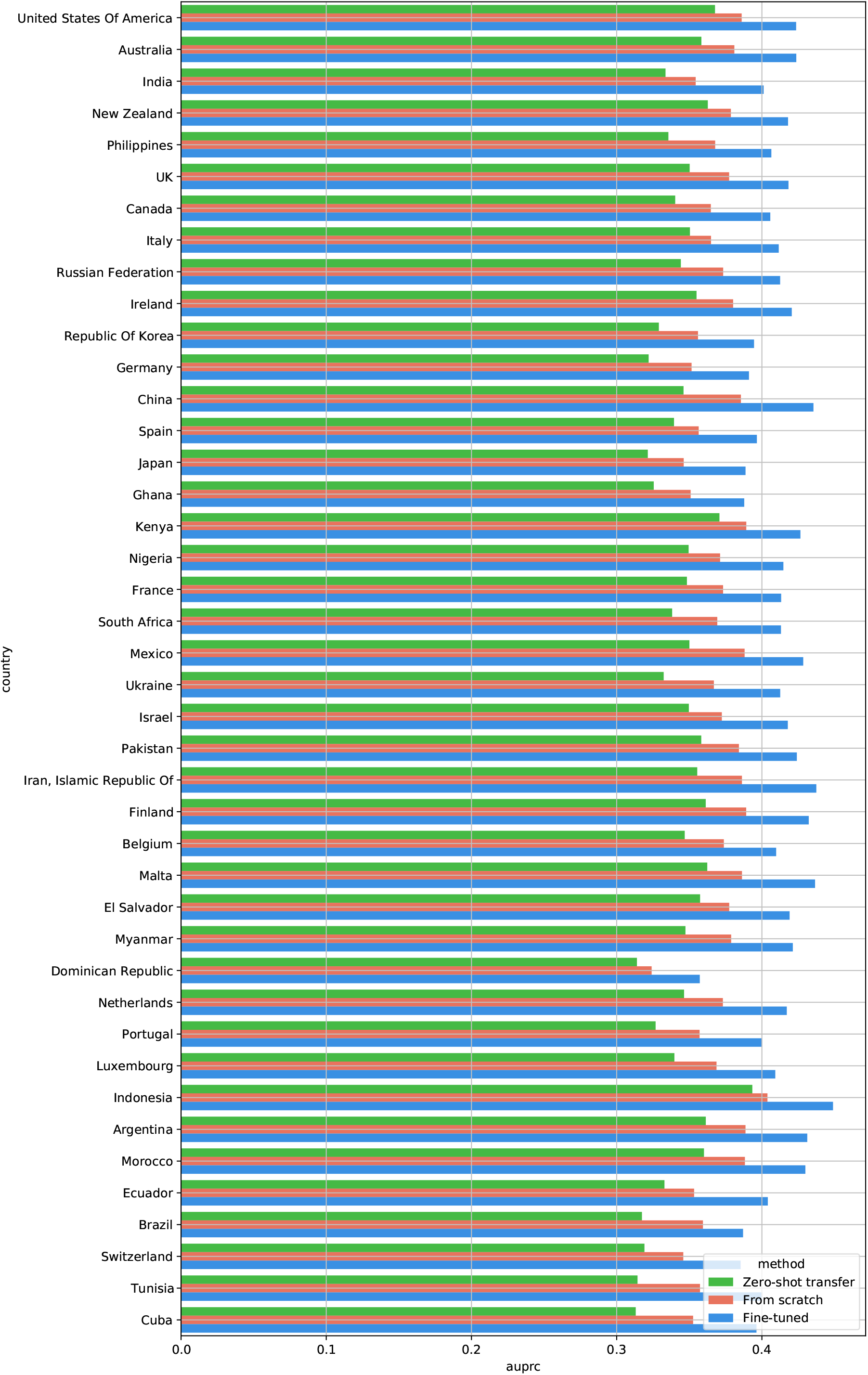
Performance of NPI prediction for each country in terms of macro AUPRC.

**Table S1:**
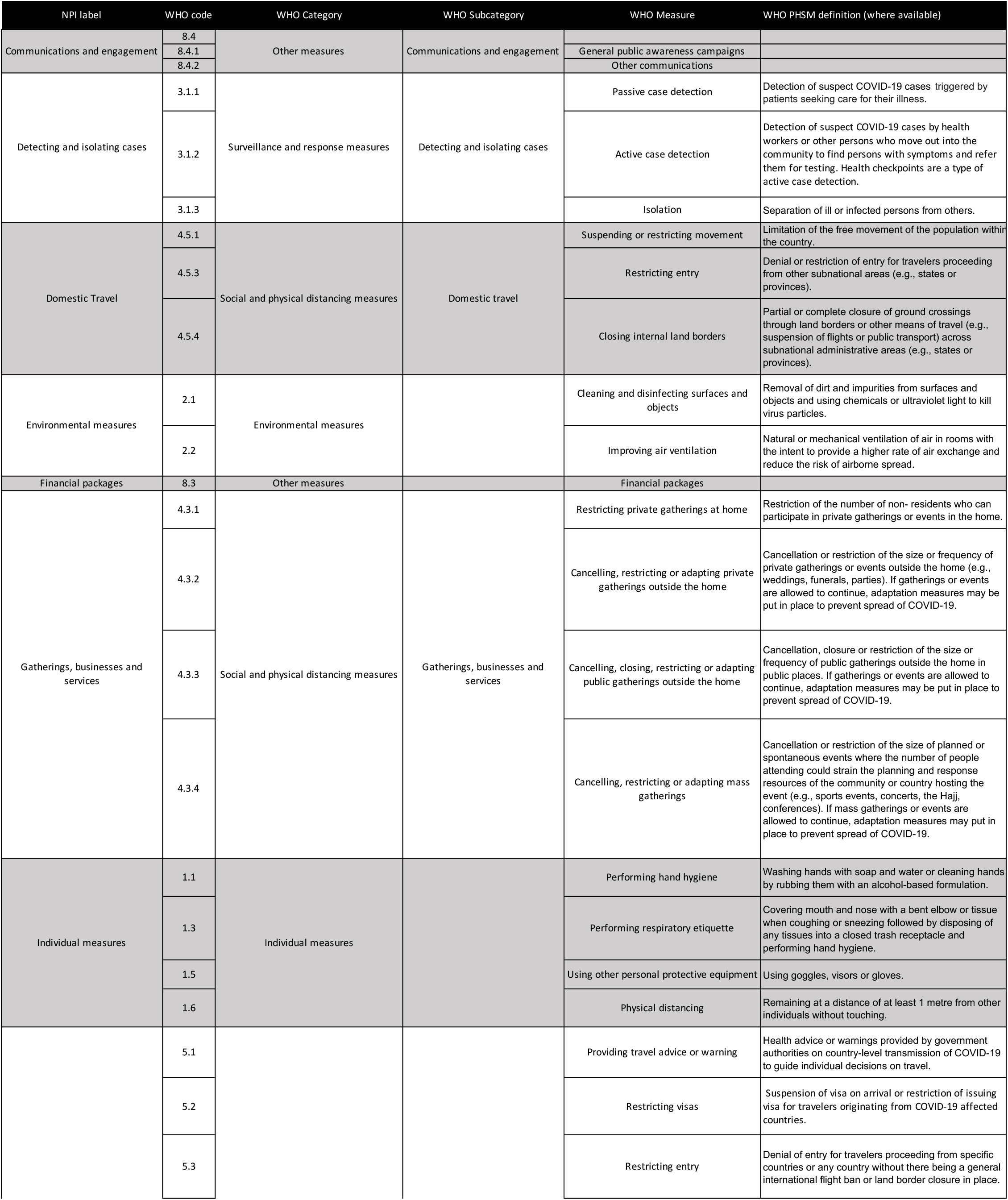
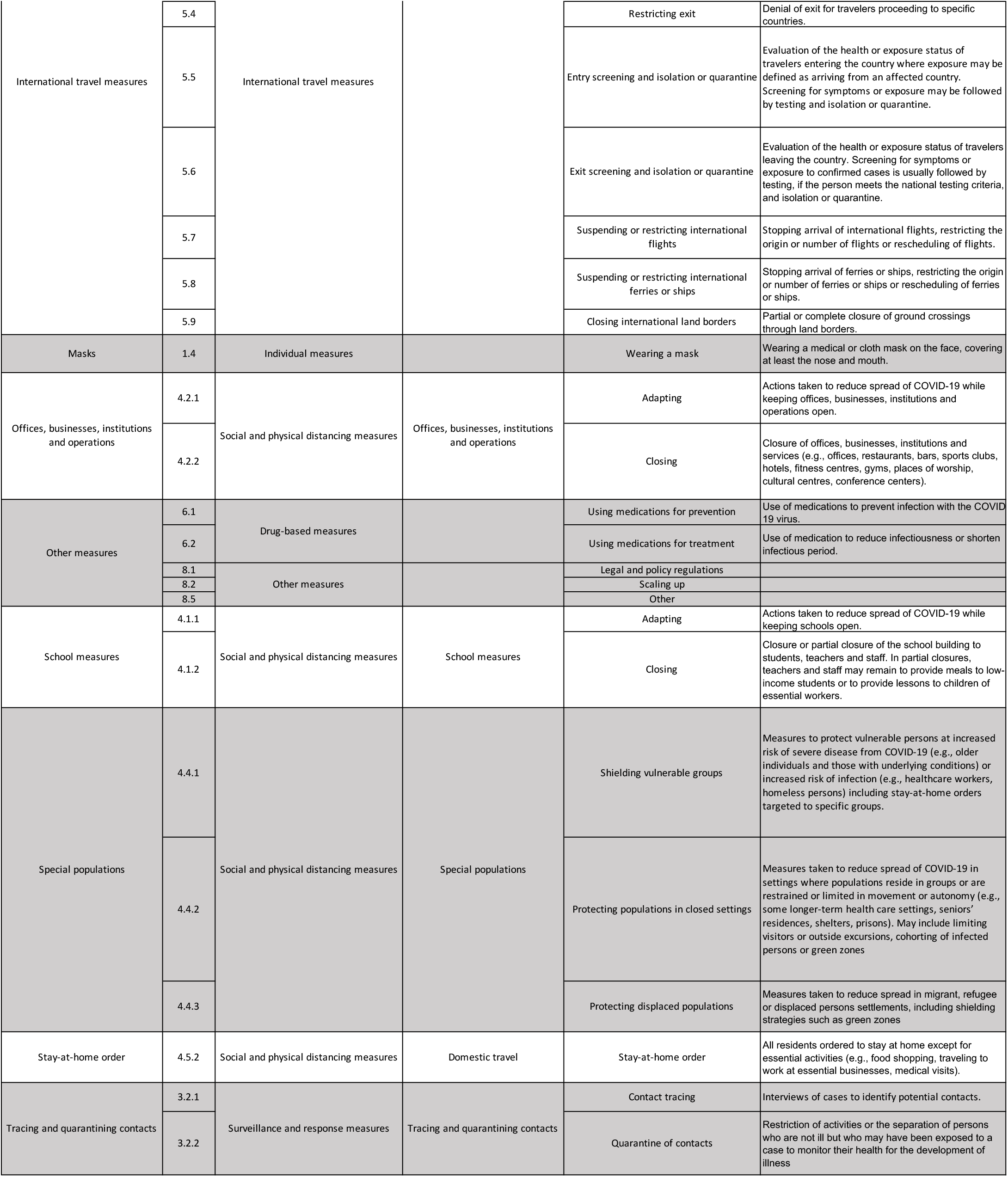
Grouping of WHO-NPI categories into 15 groups, which are treated as labels in our application of NPI predictions.

